# Aspirin and NSAID use and the risk of COVID-19

**DOI:** 10.1101/2021.04.28.21256261

**Authors:** David A. Drew, Chuan-Guo Guo, Karla A. Lee, Long H. Nguyen, Amit D. Joshi, Chun-Han Lo, Wenjie Ma, Raaj S. Mehta, Sohee Kwon, Christina M. Astley, Mingyang Song, Richard Davies, Joan Capdevila, Mary Ni Lochlainn, Carole H. Sudre, Mark S. Graham, Thomas Varsavsky, Maria F. Gomez, Beatrice Kennedy, Hugo Fitipaldi, Jonathan Wolf, Tim D. Spector, Sebastien Ourselin, Claire J. Steves, Andrew T. Chan

## Abstract

Early reports raised concern that use of non-steroidal anti-inflammatory drugs (NSAIDs) may increase risk of severe acute respiratory syndrome coronavirus 2 (SARS-CoV-2) disease (COVID-19). Users of the COVID Symptom Study smartphone application reported use of aspirin and other NSAIDs between March 24 and May 8, 2020. Users were queried daily about symptoms, COVID-19 testing, and healthcare seeking behavior. Cox proportional hazards regression was used to determine the risk of COVID-19 among according to aspirin or non-aspirin NSAID users. Among 2,736,091 individuals in the U.S., U.K., and Sweden, we documented 8,966 incident reports of a positive COVID-19 test over 60,817,043 person-days of follow-up. Compared to non-users and after stratifying by age, sex, country, day of study entry, and race/ethnicity, non-aspirin NSAID use was associated with a modest risk for testing COVID-19 positive (HR 1.23 [1.09, 1.32]), but no significant association was observed among aspirin users (HR 1.13 [0.92, 1.38]). After adjustment for lifestyle factors, comorbidities and baseline symptoms, any NSAID use was not associated with risk (HR 1.02 [0.94, 1.10]). Results were similar for those seeking healthcare for COVID-19 and were not substantially different according to lifestyle and sociodemographic factors or after accounting for propensity to receive testing. Our results do not support an association of NSAID use, including aspirin, with COVID-19 infection. Previous reports of a potential association may be due to higher rates of comorbidities or use of NSAIDs to treat symptoms associated with COVID-19.

**One Sentence Summary:** NSAID use is not associated with COVID-19 risk.

## Introduction

As the severe acute respiratory syndrome coronavirus 2 (SARS-CoV-2) disease (COVID-19) swept the globe, anecdotal reports of a potential exacerbating effect of non-steroidal anti-inflammatory drugs (NSAIDs), including ibuprofen and aspirin, on COVID-19 emerged, leading to some warnings against the use of NSAIDs(*1-4*). Concern was raised that ibuprofen, in particular, may increase angiotensin-converting enzyme 2 (ACE2) receptor expression(*5*), which may facilitate viral entry into host cells(*6-9*).

The association of medications with disease risk is often confounded by the indication for use or other comorbidities. For example, recent studies have shown that angiotensin converting enzyme inhibitors (ACEis), which were also hypothesized to predispose to risk of severe COVID-19 due to upregulation of the ACE2 pathway, have shown no increase in risk after accounting for hypertension or other cardiovascular risk factors(*10-13*) Similarly, NSAIDs are often taken by older adults for comorbid conditions that, in turn, are associated with COVID-19. In particular, aspirin is used in the primary and secondary prevention of cardiovascular disease and cancer.(*14*) NSAIDs are also often used to treat symptoms associated with COVID-19, including fever, myalgias, and sore throat. Thus, we prospectively surveyed individuals in the United States, United Kingdom, and Sweden using the COVID Symptom Study app about use of NSAIDs, risk factors for COVID-19, and disease symptoms to investigate if these agents were associated with incident COVID-19 outcomes.

## Results

### Association of use of any NSAID and COVID-19

Since March 26, we enrolled 2,736,091 study participants in the COVID Symptom Study and documented 8,966 reports of a positive COVID-19 test over 60,817,043 person-days of follow-up across the three countries. The baseline characteristics of the study cohort appear as **Table 1**. Regular NSAID users were older, were more likely to be obese (BMI ≥30) and have comorbidities including diabetes, heart, kidney, lung diseases, or cancer and report taking other medications. Compared to non-users, we observed a modest association for any NSAID users with COVID-19 (stratified hazard ratio [HR] = 1.22; 95% confidence interval [CI]: 1.13, 1.32) when stratifying for age, sex, race/ethnicity, date of study entry, and country (**Table 2a**). After additional multivariable adjustment for non-NSAID medication use and comorbidities, including diabetes, kidney, heart, and lung disease and cancer, as well as risk factors for SARS CoV-2 exposure (community or occupational contact), the comorbidity-adjusted hazard ratio was attenuated to 1.15 (95% CI: 1.06, 1.24). After further adjustment for symptoms reported at baseline, the comorbidity and symptom-adjusted HR was further attenuated to 1.02 (95% CI: 0.94, 1.10).

**Table 1.** Baseline characteristics of entire study cohort.

|  | Non-NSAID users<br>(n=2,4947,80) | Any NSAID users<br>(n=241,311) |
| --- | --- | --- |
| **Country, %** |  |  |
| United States | 5.1 | 19.4 |
| United Kingdom | 91.8 | 76.1 |
| Sweden | 3.1 | 4.5 |
| **Age (years)** | 44.0 [32.0, 57.0] | 55.00 [43.0, 67.0] |
| <25 | 12.4 | 4.1 |
| 25-34 | 18.6 | 9.7 |
| 35-44 | 20.8 | 14.3 |
| 45-54 | 19.5 | 19.9 |
| 55-64 | 16.2 | 21.6 |
| ≥65 | 12.5 | 30.4 |
|  | 0.4 | 0 |
| **Male sex, %** | 37.9 | 37.2 |
| **Race/Ethnicity, %** |  |  |
| White, non-Hispanic | 62.1 | 60.9 |
| Other | 3.6 | 3.0 |
| Prefer not to say | 0.3 | 0.3 |
| Missing | 34.1 | 35.9 |
| **BMI (kg/m<sup>2</sup>)** | 25.16 [22.32, 28.84] | 27.40 [24.11, 31.89] |
| 17-18.4 | 44.0 | 30.2 |
| 18.5-24.9 | 4.6 | 2.0 |
| 25-29.9 | 31.4 | 34.0 |
| ≥30 | 19.9 | 33.8 |
| **Comorbidities, %** |  |  |
| Diabetes | 2.8 | 8.8 |
| Heart Disease | 1.8 | 11.8 |
| Lung Disease | 11.7 | 16.7 |
| Kidney Disease | 0.6 | 1.6 |
| Cancer | 0.6 | 2.1 |
| **Medication usage, %** |  |  |
| Immunosuppressants | 2.8 | 8.2 |
| Chemo/Immunotherapy | 0.2 | 0.6 |
| Blood pressure medications | 5.6 | 23.0 |
| **Current smoker, %** | 4.1 | 3.8 |
| **Limited mobility, %<sup>†</sup>** | 4.0 | 14.8 |
| **Mobility aid, %<sup>‡</sup>** | 1.4 | 8.0 |
| **Frontline healthcare worker, %** | 5.0 | 6.1 |
| **Contact with suspected or documented COVID-19 cases, %** | 6.6 | 8.1 |
| Abbreviations: BMI (body mass index), kg (kilogram), m (meter), NSAIDs (non-steroidal anti-inflammatory drugs), S.E. (Sweden).<br>U.S. (United States), U.K. (United Kingdom)<br>Median [IQR] is presented for continuous variables.<br>Frequencies and proportions are calculated based on the total number of participants with available data.<br>History of cancer, aspirin use, and smoking status have been queried since launch in the U.S. and since 3/29/2020 in the U.K. Race and ethnicity questions were queried as of 4/17/2020 and percentages reflect the distribution among the 1,799,672 that provided data.<br><sup>†</sup> Asked as “In general, do you have any health problems that require you to stay at home?”<br><sup>‡</sup> Asked as “Do you regularly use a stick, walking frame or wheelchair to get about?” |  |  |

**Table 2.** Risk of testing positive for COVID-19 according to aspirin or NSAID use.

|  |  |  | Hazard Ratio (95% CI) |  |  |
| --- | --- | --- | --- | --- | --- |
|  | No. with Event/ Person-time (days) | 30-day incidence | Multivariable Stratification | Comorbidity-adjusted | Comorbidity and Symptom-adjusted |
| **<u>Any NSAID Use</u>** |  |  |  |  |  |
| **<u>Positive COVID-19 testing</u>** |  |  |  |  |  |
| Non users | 8191/56112463 | 0.44% | 1.0 (ref.) | 1.0 (ref.) | 1.0 (ref.) |
| Any NSAID users | 775/4704580 | 0.49% | 1.22 (1.13, 1.32) | 1.15 (1.06, 1.24) | 1.02 (0.94, 1.10) |
| **<u>Positive COVID-19 and was seen in a hospital/clinic</u>** |  |  |  |  |  |
| Non users | 1316/56112463 | 0.07% | 1.0 (ref.) | 1.0 (ref.) | 1.0 (ref.) |
| Any NSAID users | 209/4704580 | 0.13% | 1.77 (1.52, 2.06) | 1.37 (1.17, 1.60) | 1.13 (0.96, 1.32) |
| **<u>Aspirin or non-aspirin NSAID Use<sup>†</sup></u>** |  |  |  |  |  |
| **<u>Positive COVID-19 testing</u>** |  |  |  |  |  |
| Non users | 3026/18423122 | 0.49% | 1.0 (ref.) | 1.0 (ref.) | 1.0 (ref.) |
| Aspirin users | 105/1037175 | 0.30% | 1.13 (0.92, 1.38) | 1.07 (0.86, 1.33) | 1.03 (0.83, 1.28) |
| Non-aspirin NSAID users | 297/1399965 | 0.64% | 1.23 (1.09, 1.39) | 1.16 (1.02, 1.31) | 0.98 (0.87, 1.11) |
| Aspirin and NSAID users | 25/172090 | 0.44% | 1.26 (0.85, 1.88) | 1.16 (0.77, 1.74) | 0.91 (0.60, 1.38) |
| **<u>Positive COVID-19 and was seen in a hospital/clinic</u>** |  |  |  |  |  |
| Non users | 490/18423122 | 0.08% | 1.0 (ref.) | 1.0 (ref.) | 1.0 (ref.) |
| Aspirin users | 23/1037175 | 0.07% | 1.06 (0.68, 1.64) | 0.82 (0.52, 1.29) | 0.78 (0.49, 1.24) |
| Non-aspirin NSAID users | 76/1399965 | 0.16% | 1.68 (1.31, 2.15) | 1.37 (1.06, 1.76) | 1.06 (0.81, 1.38) |
| Aspirin and NSAID users | 11/172090 | 0.19% | 2.35 (1.27, 4.35) | 1.64 (0.88, 3.06) | 0.99 (0.50, 1.93) |
| Abbreviations: CI (confidence interval)<br><sup>†</sup>This analysis excludes anyone who responded prior to March 30<sup>th</sup> where aspirin use was not specifically queried separately from other NSAID use at baseline.<br>Multivariable Stratification model: Cox proportional hazards model stratified by 5-year age group, sex, race, calendar date at study entry, and country.<br>Comorbidity-adjusted model: Cox proportional hazards model stratified by factors in multivariable stratification model plus further adjustment for history of diabetes, heart disease, lung disease, kidney disease, cancer, limited mobility, mobility aid, blood pressure medications, chemotherapy/immunotherapy, immunosuppressants, and current smoker status (each yes/no), and body mass index (17-19.9, 20-24.9, 25-29.9, and ≥30 kg/m<sup>2</sup>).<br>Comorbidity and Symptom-adjusted model: Cox proportional hazards model adjusted for all factors included in the comorbidity-adjusted model with further adjustment for symptoms at baseline. |  |  |  |  |  |

### Association of any NSAID use with treatment seeking

Because early reports suggested that NSAID use may be associated with more severe cases of COVID-19, we next investigated whether there was an association between NSAID use and COVID-19 cases seeking treatment in a hospital or clinic. Stratified models demonstrated an increased risk of seeking treatment for COVID-19 (HR=1.77; 95%CI 1.52, 2.06) that was attenuated after adjusting for comorbidities and symptoms which may have led using NSAIDs at baseline (comorbidity and symptom-adjusted HR=1.13; 0.96, 1.32). Among the full cohort, we also observed no clear associations of any NSAID use with risk of COVID-19 compared to no NSAID use according to subgroups of demographic, lifestyle, and comorbidity factors (**Figure 1**) after accounting for multiple testing.

**Figure 1.**
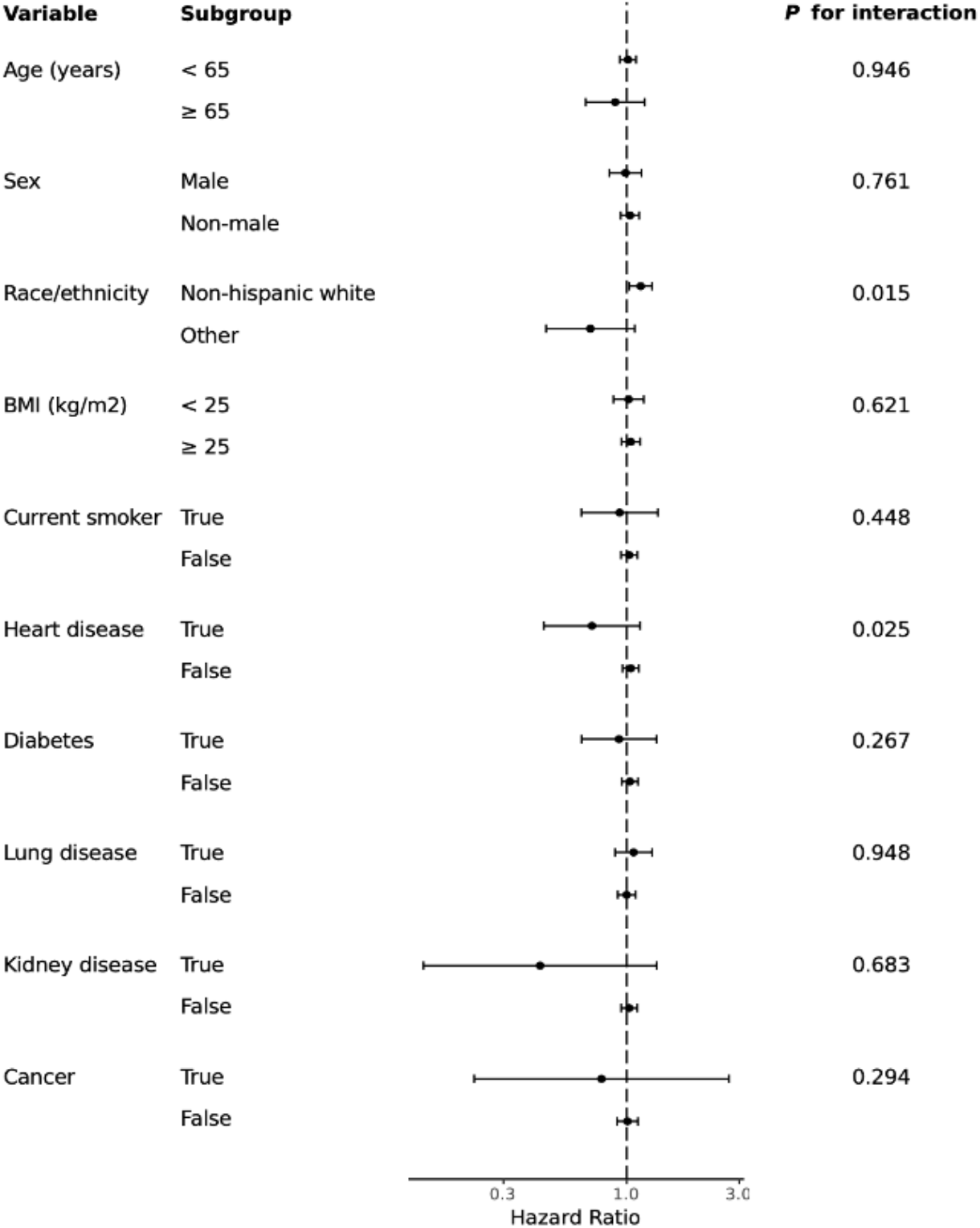
Association of NSAID use with COVID-19 according to strata of demographic, lifestyle, and comorbid factors.

### Association with COVID-19 risk according to aspirin or non-aspirin NSAID use

Within the sub-cohort of individuals who were asked separately about aspirin and other NSAID use (**Supplemental Table 1, Materials and Methods**), we also found that non-aspirin NSAID use was not associated with COVID-19 after adjustment for comorbidities, risk factors and COVID-19 symptoms at baseline which may confound use of other NSAIDs (**Table 2b**). Compared with non-users of any aspirin or NSAIDs, aspirin use was not significantly associated with risk of testing positive for COVID-19 in any models (comorbidity and symptom-adjusted HR = 1.03; 95% CI: 0.83, 1.28). Further, it appeared any potential association between NSAIDs and seeking treatment for COVID-19 was restricted to non-aspirin NSAID users with no apparent association among aspirin users. Moreover, no significant association was observed after adjusting for comorbidities and symptoms.

Overall, users of other NSAIDs reported a greater prevalence of symptoms associated with COVID-19, including those that may prompt individuals to use non-aspirin NSAIDs. The most frequently observed symptoms across the cohort were fatigue, headache, shortness of breath, and cough; however, headache, myalgias (pain), and fever appeared enriched among NSAID users, especially non-aspirin NSAID users (**Supplemental Table 2**). Individuals using aspirin reported fewer symptoms overall (17.3% reporting at least one symptom) compared to both non-users (22%) and those reporting use of other NSAIDs (34.2%) (**Supplemental Table 2**). We considered the possibility that NSAID use for treatment of symptoms and aspirin use for chronic disease prevention among those with underlying health conditions could be associated with a greater likelihood of receiving and reporting testing, potentially inducing a selection bias. However, a multivariable-adjusted IPW Cox proportional hazards model weighted by predictors of testing demonstrated no further effect on the fully adjusted association of NSAIDs with COVID-19 incidence (IPW-adjusted HR = 1.02; 95%CI: 0.93, 1.12). Further, we restricted the analysis to only those receiving testing and found no association for any NSAIDs with risk of COVID-19 in any model (comorbidity and symptom-adjusted HR=0.93; 95% CI: 0.86, 1.01) Taken together, these additional analyses confirm that NSAID use does not appear to increase the risk of testing positive for COVID-19.

## Discussion

In this prospective analysis of more than 2.7 million users of a real-time symptom tracker smartphone app, NSAID use, including use of aspirin, was not associated with a higher risk of testing positive for COVID-19. Any initial association with NSAID use is attenuated after adjustment for important confounders such as lifestyle, demographic, and comorbidity factors. There was also no observed risk of seeking or receiving treatment for COVID-19 and this association was not impacted by the propensity to receive testing compared to individuals reporting no NSAID use.

These analyses indicate that initial anecdotal reports of a potentially harmful association of NSAID use with COVID-19 incidence due to hypothesized upregulation of ACE2 receptor expression were likely related to confounding. Predictors of COVID-19 severity (i.e. heart disease) and symptoms related to acute illness are both indications for which aspirin or non-aspirin NSAID use, respectively, may be recommended, which may have contributed to a positive association. Our results after adjusting for comorbidities and symptoms demonstrate that any putative risk of COVID-19 associated with reported NSAID use is likely due, in part, to these factors driving use of aspirin or NSAIDs. Baseline use of NSAIDs was nominally associated with COVID-19 risk and was non-significant among those cases tested or seeking treatment. This may suggest that individuals feeling unwell with COVID-19 symptoms (e.g. fatigue, aches/pains, fever or chills) may be more likely to use NSAIDs, particularly non-aspirin NSAIDS, for symptom management prior to seeking testing or treatment. As outlined by the FDA’s guidance(*3*), labels on over-the-counter NSAIDs warn that “the pharmacological activity of NSAIDs in reducing inflammation, and possibly fever, may diminish the utility of diagnostic signs in detecting infections.” Thus, individuals using NSAIDs for early symptoms may have been more likely to present to hospitals and clinics later in the disease course leading to anecdotal reports of higher disease severity associated with NSAID use. Indeed, we observed that reported symptom burden was higher among non-aspirin NSAID users. However, we found no increased risk of testing positive for COVID-19 when we weighted risk estimates for the propensity to receive a test or restricted our analyses to models to those who received testing. Furthermore, there was no association of aspirin, an agent which is not generally used for management of symptoms, and risk of testing positive.

These findings have important implications for aspirin use during the pandemic. Aspirin, unlike other NSAIDs (e.g. ibuprofen, naproxen), is more likely to be used chronically for its established anti-platelet effects via inhibition of prostaglandin synthase 1 (PTGS1 or cyclooxygenase [COX-1])(*15, 16*) and recommended use in primary or secondary prevention of cardiovascular/thrombotic events and colorectal cancer(*17*). Moreover, the population most likely to use aspirin are those with comorbidities such as cardiovascular disease or diabetes, which have been associated with more severe COVID-19 outcomes.(*6-8*) Our data reassures patients and providers that decision-making regarding aspirin use need not incorporate concerns related to risk for COVID-19 infection. Moreover, given emerging indications that venous thromboembolism is a prevalent complication in COVID-19(*18*) and suggestions that early anticoagulant intervention may improve prognosis(*19*), unnecessary cessation of maintenance aspirin could be particularly harmful for patients due to the potential for rebound hypercoagulability. Further studies are needed to examine a potential role for aspirin in the prevention of such complications with severe COVID-19 as well as if outcomes among those with established infection differ according to NSAID use, independent of comorbidities.

Our study has many strengths, including prospective collection of key relevant lifestyle and demographic factors, comorbidities, and symptom data among a large study population with sufficient longitudinal follow-up for clinical trajectories and COVID-19 infections during a critical phase of the pandemic. However, there are several limitations to our study. The nature of data collection relies on self-report and community testing practices. While we have accounted for multiple potential sources of potential confounding, there may be residual bias according to unmeasured confounders (e.g. access to healthcare, health-conscious behavior), as well as measurement bias (e.g. recall-bias for self-reports, NSAID use in response to COVID-19 infection), or selection bias (e.g. differential app participation or testing by NSAID use). To address these concerns, we adjusted for comorbidities for which aspirin use is an indication (e.g. heart disease) or baseline symptoms likely to prompt NSAID use (e.g. fever or headache). We also conducted sensitivity analyses in which we adjusted for the propensity to receive testing using IPW and restricted to those that received testing. Last, our approach through voluntary reporting is unlikely to be able to capture the effects of NSAIDs in relation to the most severe COVID-19 symptoms and complications.

In conclusion, we find no evidence to support an increased risk for COVID-19 in those that regularly use aspirin or non-aspirin NSAIDs. These findings may offer reassurance to patients and providers regarding the safety of continuing use of aspirin for prevention of cardiovascular disease or cancer and the use of NSAIDs for treatment of chronic pain or other inflammatory conditions during the COVID-19 pandemic.

## Materials and Methods

### The COVID Symptom Study smartphone application

The COVID Symptom Study smartphone application (“app”) is a freely available program developed by Zoe Global Ltd. with scientific input from researchers and clinicians at Massachusetts General Hospital and King’s College London that offers participants the ability to report a range of baseline demographic information and comorbidities upon first use, as previously described.(*20*) Participants are encouraged to use the app daily, even when asymptomatic, to allow for the longitudinal, prospective collection of concomitant symptoms, COVID-19 testing results, treatment and outcomes. The app was launched in the UK (March 24, 2020), U.S. (March 29, 2020) and Sweden (April 29, 2020). The analyses were performed on data provided by all participants enrolled in the study through May 8th, 2020. Participants were recruited from the general public through media, social media, and traditional study recruitment outreach. Participants provided informed consent to the use of app data for research purposes and agreed to privacy policies and terms of use. This research study was approved by the Partners Human Research Committee (IRB 2020P000909), King’s College London Ethics Committee (REMAS ID 18210, Review Reference LRS-19/20-18210), and the central ethics committee in Sweden (DNR 2020-01803).

### Assessment of NSAID use, risk factors, symptoms, testing, and treatment

On initiation of the study on March 24, 2020, participants were asked to provide demographic factors and answered separate questions about possible risk factors for COVID-19, including comorbidities and medication use, including NSAIDs, at baseline. Starting on March 30^th^ (coinciding with launch of the app in the U.S.) we began asking new users separate questions to distinguish regular aspirin use from other non-aspirin NSAIDS. After enrollment, participants are prompted daily to report if they felt physically normal, and if not, their symptoms, including those possibly related to COVID-19 (i.e. presence of fever, persistent cough, fatigue, loss of smell/taste, and diarrhea, among others), as well as health care seeking behavior (i.e. whether they had been to the hospital and/or sought and received clinical treatment, including supplemental oxygen, invasive ventilation, intravenous fluids, or inhalers). Participants were also asked if they had been tested for COVID-19 (yes/no), and if yes, the results (none, negative, waiting for results, or positive).

### Statistical analysis

For the main analysis, we included all individuals except those that had reported a positive COVID-19 test within 24 hours of enrolling in the study. We employed Cox proportional hazards regression to examine the association between regular use of any NSAID and risk of a positive COVID-19 test. The initial models (referred to as “multivariable stratified”) were stratified by age, sex, date, country, and race/ethnicity. Subsequent models (referred to as “comorbidity-adjusted”) additionally adjusted for covariates that were selected *a priori* based on putative risk factors and included self-reported height and weight calculated body mass index (<18.5, 18.5-24.9, 25-29.9, and ≥30 kg/m2), history of diabetes, heart disease, lung disease, kidney disease, current smoking status (each yes/no), limited mobility (homebound or walks with assistance of a cane or walker). Missing categorical data were included as a missing indicator. Because NSAID use, particularly non-aspirin NSAID use at baseline, may be confounded by COVID symptoms (i.e. fever, headache, aches/pain) that lead to their use, we additionally adjusted analyses for any baseline symptoms that were reported (referred to as “symptom adjusted”). To understand the differences associated with regular aspirin use versus NSAID use, we also limited the analysis to the sub-cohort that was asked separately about these beginning on March 30. In sensitivity analyses, we limited the analysis to individuals who received testing. Using inverse probability weighting (IPW) for receiving a test based on age, country of residence, contact with individuals with suspected or documented COVID-19, being a healthcare worker, and reporting symptoms at baseline, we also conducted an IPW-adjusted Cox model to account for the propensity to receive a test.(*21*) In a secondary analysis, we examined the association of NSAIDs with COVID-19 requiring hospitalization or treatment as a proxy for disease severity. In addition, we stratified models according to comorbidities, lifestyle and sociodemographic factors to assess whether differences for the association between NSAIDs and COVID-19 or an interaction between factors and NSAIDs through inclusion of a multiplicative interaction term in the model was present among subgroups of the study population. Two-sided *p-values* <0.05 were considered statistically significant for the primary and secondary analyses. Interaction tests in stratified analyses were considered significant at p<0.005 using Bonferroni multiple hypothesis adjustment (α=0.05/10 stratification factors = 0.005). All analyses were performed using R 3.6.1 (Vienna, Austria).

## Data Availability

Data collected in the app is being shared with other health researchers through the NHS-funded Health Data Research U.K. (HDRUK)/SAIL consortium, housed in the U.K. Secure Research Platform (UKSeRP) in Swansea. Anonymized data is available to be shared with bonafide researchers HDRUK according to their protocols (https://healthdatagateway.org/detail/9b604483-9cdc-41b2-b82c-14ee3dd705f6). U.S. investigators are encouraged to coordinate data requests through the COPE Consortium (www.monganinstitute.org/cope-consortium). Data updates can be found on https://covid.joinzoe.com.

## Supplementary Materials

**Supplemental Table 1.** Baseline characteristics according to use of aspirin, non-aspirin NSAIDs, or Both (Cohort restricted to 3/30 baseline)

|  | Non users<br>(n=1,255,781) | Aspirin users<br>(n=653,86) | Non-aspirin NSAID users<br>(n=93,867) | Aspirin and NSAID users<br>(n=11,293) |
| --- | --- | --- | --- | --- |
| **Country, %** |  |  |  |  |
| U.S. | 10.0 | 30.1 | 22.8 | 46.0 |
| U.K. | 83.8 | 64.5 | 70.6 | 44.8 |
| S.E. | 6.1 | 5.4 | 6.6 | 9.2 |
| **Age (years)** | 47.0 [32.0, 60.0] | 68.0 [60.0, 74.0] | 52.0 [41.0, 63.0] | 65.00 [54.0, 72.0] |
| <25 | 16.2 | 0.6 | 4.7 | 2.6 |
| 25-34 | 12.7 | 1.6 | 9.6 | 4.1 |
| 35-44 | 16.3 | 3.1 | 16.5 | 6.2 |
| 45-54 | 19.3 | 9.1 | 24.4 | 12.7 |
| 55-64 | 18.1 | 24.1 | 22.8 | 24.1 |
| ≥65 | 17.4 | 61.5 | 22.1 | 50.3 |
| **Male sex, %** | 43.9 | 58.8 | 31.5 | 44.6 |
| **Race/Ethnicity, %** |  |  |  |  |
| White, non-Hispanic | 72.5 | 68.7 | 63.8 | 55.0 |
| Other | 4.3 | 4.0 | 3.0 | 3.3 |
| Prefer not to say | 0.3 | 0.3 | 0.3 | 0.3 |
| **BMI (kg/m<sup>2</sup>)** | 25.1 [22.1, 28.8] | 27.1 [24.2, 30.7] | 27.6 [24.1, 32.3] | 28.3 [24.9, 32.9] |
| 17-18.4 | 41.8 | 30.2 | 29.5 | 24.1 |
| 18.5-24.9 | 7.0 | 1.9 | 2.2 | 2.1 |
| 25-29.9 | 31.3 | 39.2 | 32.6 | 34.3 |
| ≥30 | 19.9 | 28.6 | 35.7 | 39.5 |
| **Comorbidities, %** |  |  |  |  |
| Diabetes | 3.4 | 17.3 | 5.6 | 16.4 |
| Heart Disease | 2.2 | 33.1 | 2.7 | 20.6 |
| Lung Disease | 11.5 | 14.6 | 17.1 | 19.1 |
| Kidney Disease | 0.7 | 3.8 | 0.9 | 2.0 |
| Cancer | 1.2 | 4.4 | 1.8 | 3.7 |
| **Medication usage, %** |  |  |  |  |
| Immunosuppressants | 3.1 | 6.9 | 8.6 | 10.8 |
| Chemo/Immunotherapy | 0.2 | 1.0 | 0.3 | 0.8 |
| Blood pressure medications | 9.6 | 49.5 | 16.7 | 46.0 |
| **Current smoker, %** | 0.1 | 0 | 0.2 | 0 |
| **Limited mobility, %†** | 6.4 | 21.0 | 14.8 | 21.4 |
| **Mobility aid, %‡** | 2.0 | 8.7 | 8.2 | 13.8 |
| **Frontline healthcare worker, %** | 6.1 | 4.1 | 8.6 | 6.8 |
| **Interaction with suspected or documented COVID-19, %** | 12.1 | 6.8 | 13.7 | 9.6 |
| Abbreviations: BMI (body mass index), kg (kilogram), m (meter), NSAIDs (non-steroidal anti-inflammatory drugs), S.E. (Sweden). U.S. (United States), U.K. (United Kingdom) |  |  |  |  |
| Median [IQR] is presented for continuous variables. |  |  |  |  |
| Frequencies and proportions are calculated based on the total number of participants with available data. |  |  |  |  |
| History of cancer, aspirin use, and smoking status have been queried since launch in the U.S. and since 3/29/2020 in the U.K. Race and ethnicity questions were queried as of 4/17/2020 with 1,085,282 participants providing data. |  |  |  |  |
| † Asked as “In general, do you have any health problems that require you to stay at home?” |  |  |  |  |
| ‡ Asked as “Do you regularly use a stick, walking frame or wheelchair to get about?” |  |  |  |  |

**Supplemental Table 2.**
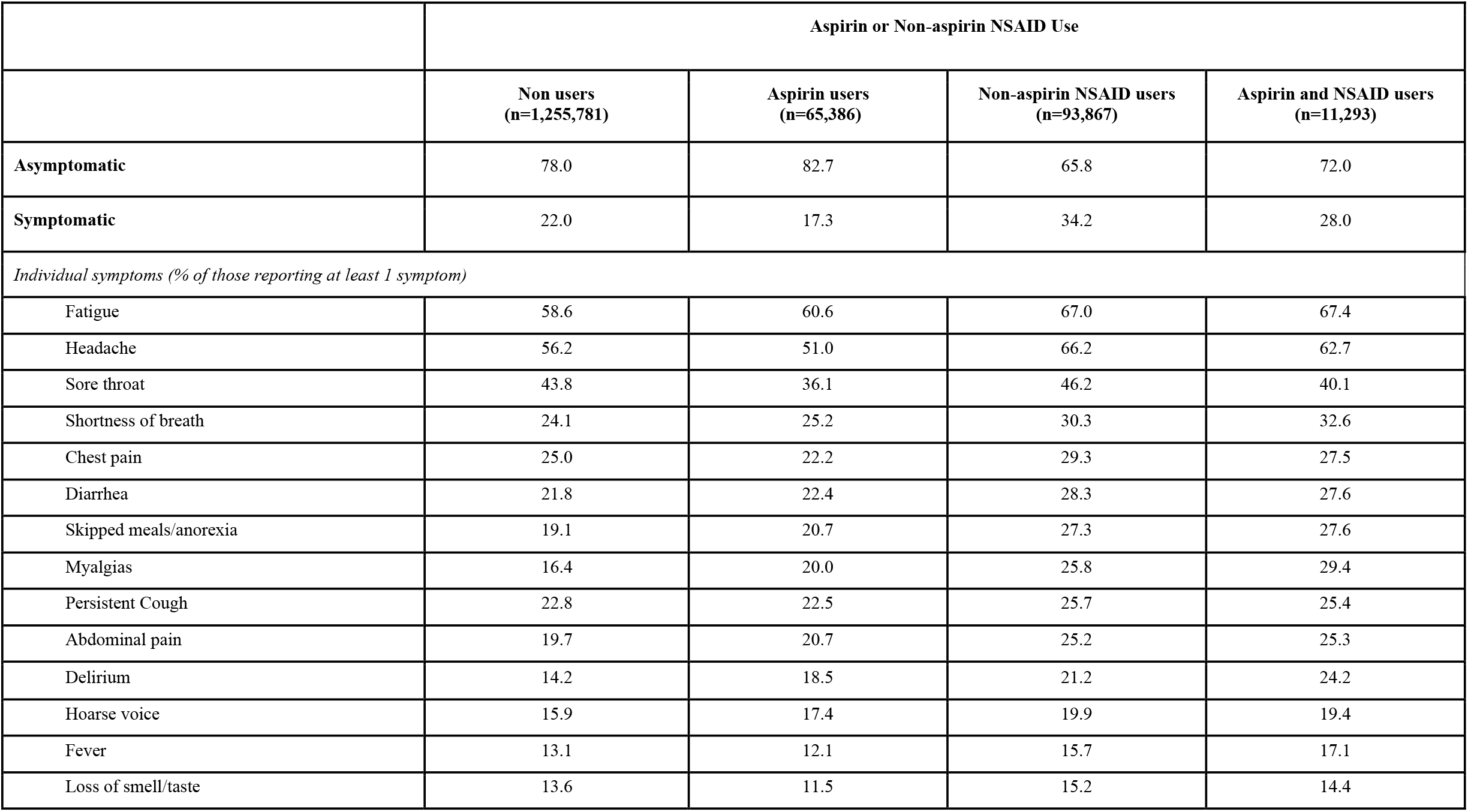
Aspirin and non-aspirin NSAIDs and symptom occurrence.

|  | Aspirin or Non-aspirin NSAID Use |  |  |  |
| --- | --- | --- | --- | --- |
|  | Non users<br>(n=1,255,781) | Aspirin users<br>(n=65,386) | Non-aspirin NSAID users<br>(n=93,867) | Aspirin and NSAID users<br>(n=11,293) |
| **Asymptomatic** | 78.0 | 82.7 | 65.8 | 72.0 |
| **Symptomatic** | 22.0 | 17.3 | 34.2 | 28.0 |
| *Individual symptoms (% of those reporting at least 1 symptom)* |  |  |  |  |
| Fatigue | 58.6 | 60.6 | 67.0 | 67.4 |
| Headache | 56.2 | 51.0 | 66.2 | 62.7 |
| Sore throat | 43.8 | 36.1 | 46.2 | 40.1 |
| Shortness of breath | 24.1 | 25.2 | 30.3 | 32.6 |
| Chest pain | 25.0 | 22.2 | 29.3 | 27.5 |
| Diarrhea | 21.8 | 22.4 | 28.3 | 27.6 |
| Skipped meals/anorexia | 19.1 | 20.7 | 27.3 | 27.6 |
| Myalgias | 16.4 | 20.0 | 25.8 | 29.4 |
| Persistent Cough | 22.8 | 22.5 | 25.7 | 25.4 |
| Abdominal pain | 19.7 | 20.7 | 25.2 | 25.3 |
| Delirium | 14.2 | 18.5 | 21.2 | 24.2 |
| Hoarse voice | 15.9 | 17.4 | 19.9 | 19.4 |
| Fever | 13.1 | 12.1 | 15.7 | 17.1 |
| Loss of smell/taste | 13.6 | 11.5 | 15.2 | 14.4 |

## Acknowledgments

We would like to thank the contributing citizen researchers who participated in the COVID Symptom Study and participants of the studies within the COronavirus Pandemic Epidemiology (COPE) Consortium.

## Author contributions

DAD, CGG, and ATC performed formal analysis. DAD, CGG, and KAL drafted the initial manuscript. All authors contributed to the conceptualization, methodology, investigation, resources, data curation, and review and editing of the manuscript. ATC supervised the study.

